# Beneficial and harmful outcomes of tocilizumab in severe COVID-19: a systematic review and meta-analysis

**DOI:** 10.1101/2020.09.05.20188912

**Authors:** Manuel Rubio-Rivas, Jose María Mora-Luján, Abelardo Montero, Narcís A. Homs, Jordi Rello, Xavier Corbella

## Abstract

**Objectives:** Pending for randomized control trials, the use of tocilizumab (TCZ) in COVID-19 remains controversial. We performed a systematic review and meta-analysis to investigate the effect on clinical outcomes of TCZ to treat severe COVID-19.

**Methods:** From 1 January to 21 August 2020, we searched PubMed (via MEDLINE), Scopus, and medRxiv repository databases for observational studies in any language reporting efficacy and safety of TCZ use in hospitalized adults with COVID-19. Independent and dually data extraction and quality assessment were performed.

**Results:** Of 57 eligible studies, 27 controlled and 30 not. The overall included patients were 8,128: 4,021 treated with TCZ, in addition to standard of care (SOC), and 4,107 only receiving SOC. The pooled mortality was lower in the TCZ-group, with a relative risk (RR) of 0.73 (95%CI 0.57-0.93; p=0.010). TCZ-treated patients were transferred to the intensive care unit (ICU) in a higher proportion, but ICU mortality was lower than in the control group. Conversely, a higher proportion of TCZ-treated patients developed secondary infections after TCZ use.

**Conclusions:** TCZ seems beneficial in preventing in-hospital mortality in severe, non-critically ill COVID-19 patients. However, patients receiving TCZ appear to be at higher risk for secondary infections, especially those admitted to ICU.

## INTRODUCTION

Since early 2020, when the SARS-CoV-2 pandemic hit the world, a variety of treatments have been suggested for COVID-19 [1-5]. However, to date, only remdesivir [4] and dexamethasone [5] have demonstrated evidence-based efficacy on randomized, controlled clinical trials (RCTs). This double strategy combining antiviral and immunomodulatory therapy is in accordance with the two pathological mechanisms that appear to coexist in the disease; the first triggered by the virus itself and the second by the cytokine storm and systemic dysregulated host-immune hyperinflammatory response [6].

While the pandemic continues to spread globally, a worrying 15% of patients continue to transit into the most severe stage of the disease, requiring hospitalization or intensive care unit (ICU) admission. This advanced clinical-stage presents as severe pulmonary injury and multi-organ failure, causing fatality in nearly half of cases, resembling complications from CAR T cell therapy [7].

Among other pro-inflammatory cytokines, IL-6 plays a part in innate immunity, but excessive production by the host facing SARS-CoV-2 is detrimental [8-10]. Accordingly, the use of immunomodulatory agents such as tocilizumab (TCZ), a monoclonal antibody to the recombinant human IL-6 receptor, was initially reported as successful among 21 patients in China [11]. Since then, an emerging number of observational studies from America and Europe have been published or registered assessing the effect of TCZ in severe COVID-19 [12-67]. In most of them, the authors report an association between earlier use of TCZ and reduced mortality; however, interpretation of these results is limited because several of them did not describe a comparison group.

Conversely, given preliminary results from the industry-sponsored Phase 3 COVACTA trial (ClinicalTrials.gov Identifier NCT04320615), the COVID-19 Treatment Guidelines Panel by the National Institute of Health, has taken a position against the use of TCZ [68]. This RCT is the first global, randomized, double-blind, placebo-controlled phase III trial investigating TCZ in hospitalized patients with severe COVID-19, but it failed to demonstrate improvement in clinical status as the primary endpoint, or several key secondary outcomes such as 4-week mortality [69].

In the current emergency, while waiting for additional data from RCTs, the fact that most institutions and physicians worldwide are still tackling COVID-19 based only on real-world reported data, prompted us the present review. Our aim was to summarize the updated results from available observational studies on the effect of TCZ on clinical outcomes in hospitalized patients with COVID-19.

## METHODS

This systematic review and meta-analysis followed the guidance of the Preferred Reporting Items for Systematic Reviews and Meta-analyses (PRISMA) statement [70]. The protocol was published in the National Institute for Health Research international register of systematic reviews (PROSPERO); registration number CRD42020204934. A clinical question under the PICO framework format (Population-Intervention-Comparison-Outcome) was created (Table 1).

**Table 1.**
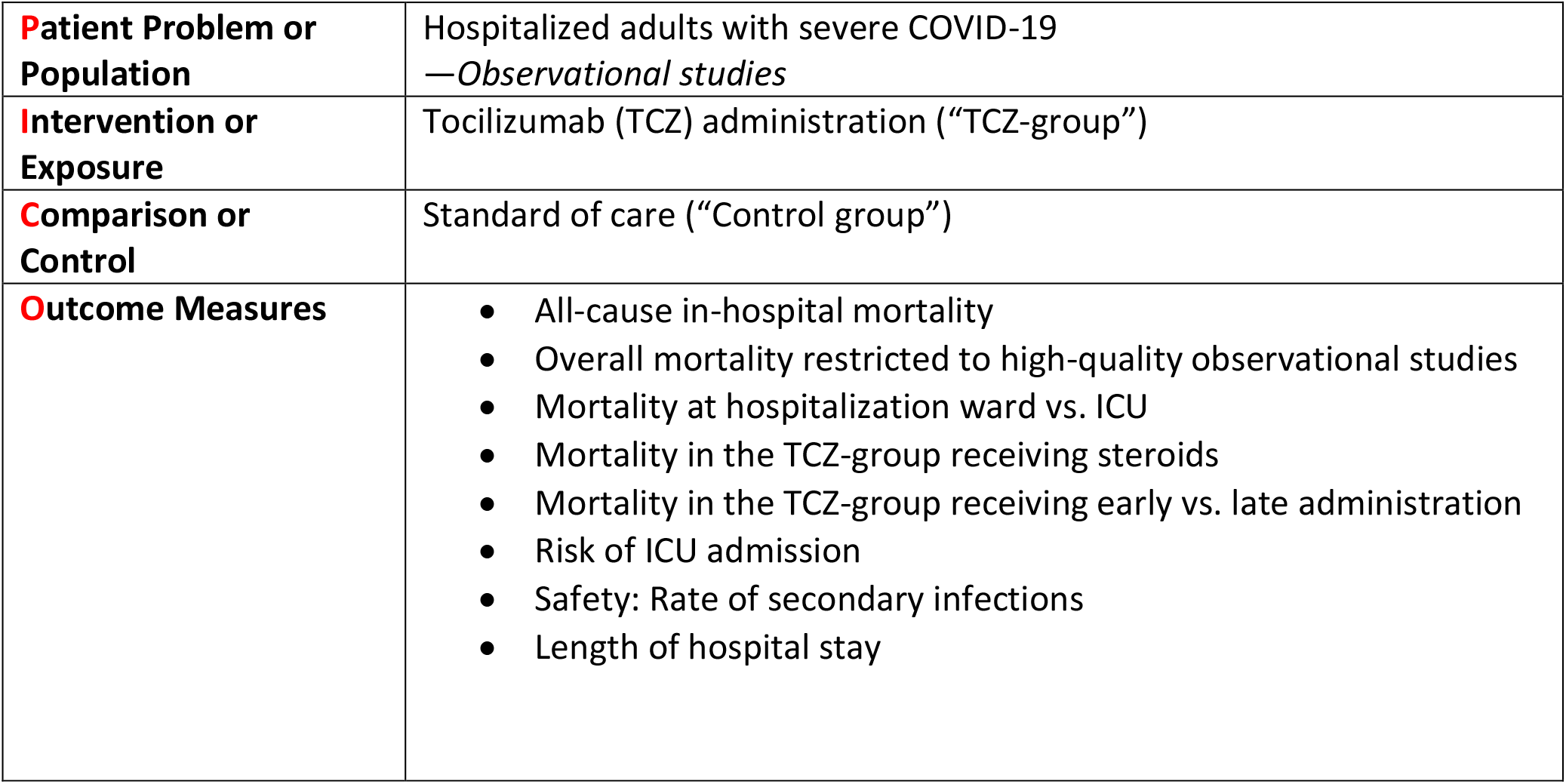
PICO framework format

|  |  |
| --- | --- |
| **Patient Problem or Population** | Hospitalized adults with severe COVID-19<br>—*Observational studies* |
| **Intervention or Exposure** | Tocilizumab (TCZ) administration (“TCZ-group”) |
| **Comparison or Control** | Standard of care (“Control group”) |
| **Outcome Measures** | • All-cause in-hospital mortality<br>• Overall mortality restricted to high-quality observational studies<br>• Mortality at hospitalization ward vs. ICU<br>• Mortality in the TCZ-group receiving steroids<br>• Mortality in the TCZ-group receiving early vs. late administration<br>• Risk of ICU admission<br>• Safety: Rate of secondary infections<br>• Length of hospital stay |

### Data Sources and Searches

The search strategy was developed by three investigators (M.R-R., J.R., X.C.), which was revised and approved by the other investigators (J.M.M-L., A.M, N.A.H.). We searched the following databases from 1 January to 21 August 2020: MEDLINE database through the PubMed search engine, Scopus, and the medRxiv repository, using the terms “COVID-19” [MesH]) AND “Tocilizumab” [MesH].

### Study Eligibility Criteria

Full-text observational studies in any language reporting beneficial or harmful outcomes from the use of TCZ in adults hospitalized with COVID-19 were included. Two investigators (M.R-R., J.M.M-L.) independently screened each record title and abstract for potential inclusion. Restriction of publication type was manually applied: secondary analyses of previously reported trials, protocols, abstracts-only and experimental studies were excluded. Potentially relevant articles were retrieved for full-text review. Two investigators (M.R-R., A.M.) read the full text of the abstracts selected. Discrepancies were resolved through discussion or by a third investigator (X.C.).

Publications were included if they met all the following criteria: 1) the study reported data on adults ≥18 years-old with COVID-19, diagnosed by polymerase chain reaction (PCR), admitted hospital-wide or in ICUs; 2) the study design was an observational investigation providing real-world original data on TCZ use in COVID-19, either intravenous or subcutaneous; and 3) the study data collection finished after 1 January 2020.

Those studies reported being “case-control studies”, in which subjects from the control group also presented COVID-19, just as those from the TCZ group, were also included in the present review. Studies focusing on a sole subgroup of patients (e.g. renal transplant recipients) were excluded. A careful revision was also performed of patients’ origins included in studies from the same country/hospital to avoid overlapping data, and only the latest and largest study was selected. The search was completed by the bibliography review of every paper selected for full-text examination.

### Data Extraction and Quality Assessment

Two investigators (M.R-R., J.M.M-L.) independently abstracted the following details: study characteristics, including setting; intervention or exposure characteristics, including medication dose and duration; patient characteristics, including the severity of disease; and outcomes, including mortality, admission to ICU, adverse events such as secondary infections, and length of hospital stay. Discrepancies were resolved by discussion in consultation with a third investigator (X.C.). Quality assessment was performed by two investigators (M.R-R., N.A.H) using the Newcastle-Ottawa Scale (NOS) for observational studies [71]. In case of disagreement, a third author (J.R.) independently determined the quality assessments.

### Data Analysis

Categorical variables were described as absolute numbers and percentages. When the number of events and the sample size was small (and followed a Poisson distribution), confidence intervals were estimated using Wilson’s method [72-74]. We carried out a meta-analysis of the pooled mortality ratio by including all comprised studies. Those studies with a control group were also meta-analyzed to assess the relative risk (RR) of mortality in TCZ-treated patients vs. those non-TCZ treated (RR of 1).

The inverse variance-weighted method was initially performed using a fixed-effects model. Heterogeneity between studies was assessed using the Q statistic. The percentage of variability between studies by the Higgins I^2^ parameter and between-study variability was measured by the Tau^2^ parameter and, when confirmed (p≤0.05), the analysis was completed by using the random-effects model [75]. Studies with 0 events in the only arm (uncontrolled studies) or both arms (controlled studies) were not included in the meta-analysis. Forest plots were depicted accordingly. Publication bias was assessed using the Egger method [76]. Statistical analysis was performed by IBM SPSS Statistics for Windows, Version 26.0. Armonk, NY: IBM Corp.

## RESULTS

A total of 781 articles were identified in our search. Of these, 81 qualified for full-text review following title and abstract screening, of which 57 [11-67] were included in the analysis. The PRISMA flow diagram is detailed in Figure 1.

**Figure 1.**
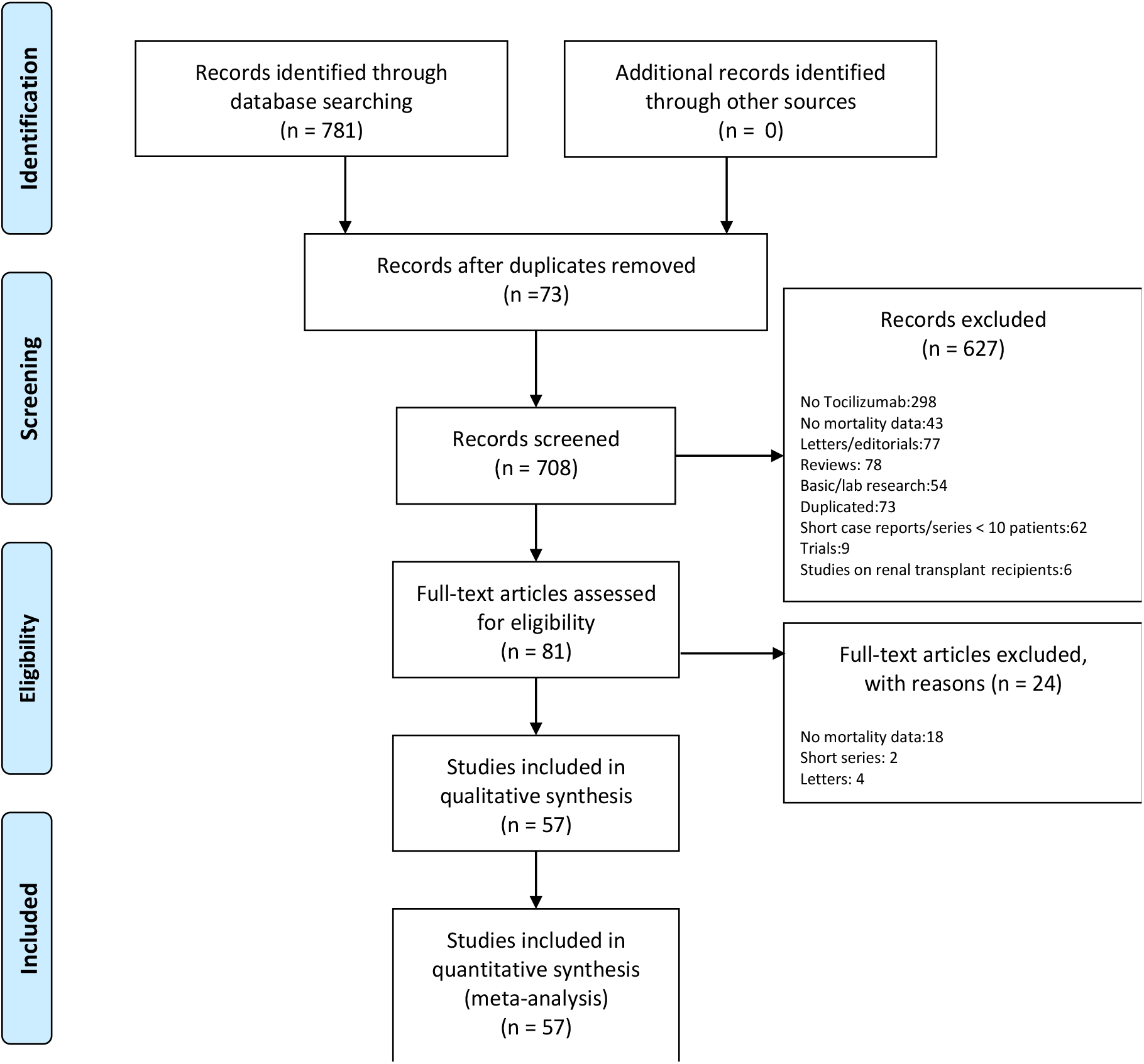
PRISMA 2009 Flow Diagram

### Study characteristics

The majority of included studies were carried out in different hospitals in high-income countries in America and Europe, such as the US [13, 18, 22, 28, 31, 34, 35, 48, 49, 53-60, 66], Italy [14, 15, 17, 19, 20, 41, 44-47, 50, 51, 63, 65], Spain [21, 26, 27, 32, 33, 39, 40, 42, 43], and France [25, 29, 30, 61, 62, 64]. A lesser number were conducted in China [11, 12]. The distribution of the studies worldwide is shown in Figure 2.

**Figure 2.**
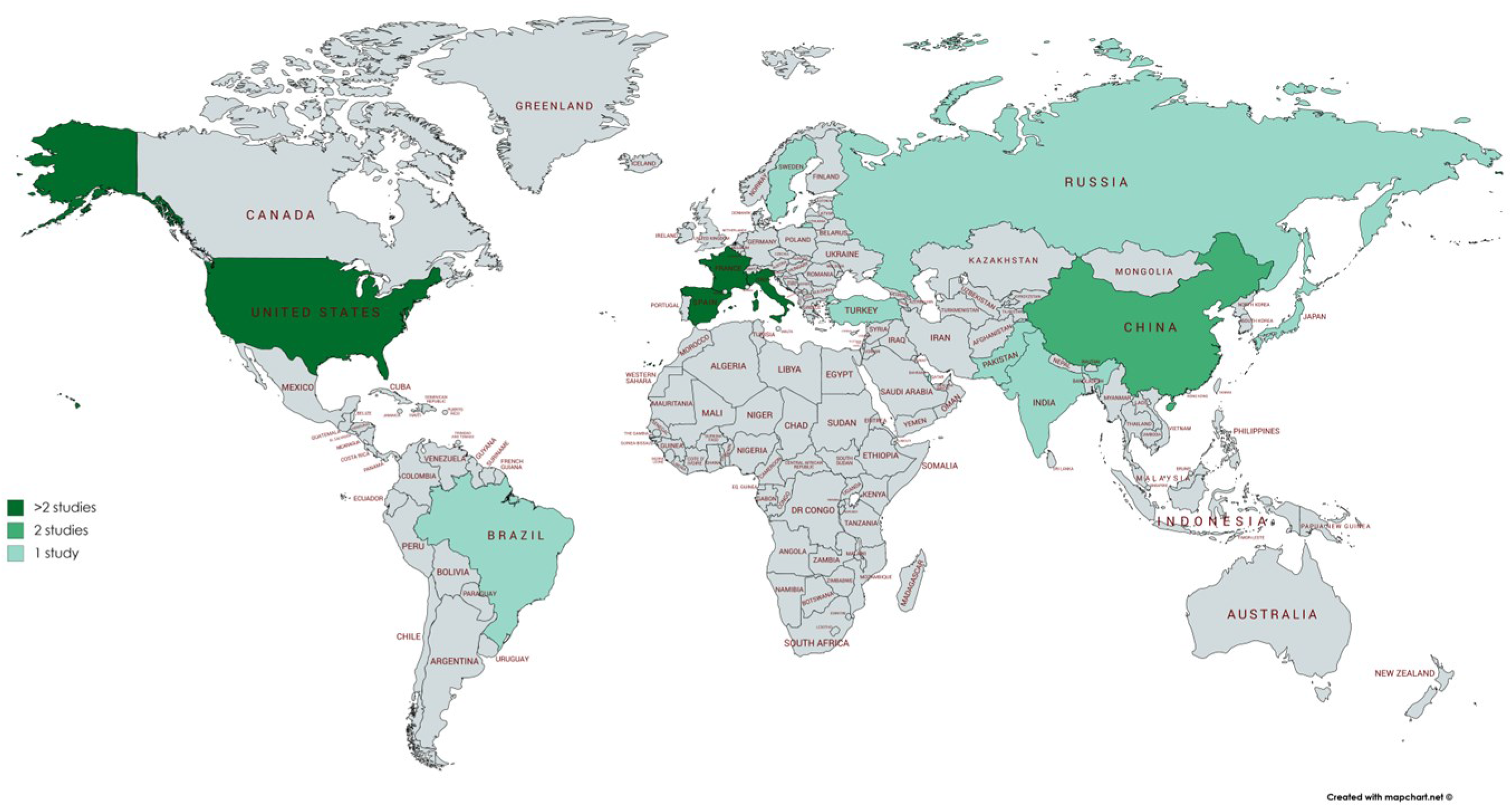
Worldwide Distribution of the Included Studies.

### Search results

Of the total of 57 studies included, 12 were prospective and 45, retrospective. In 30 of these cohort investigations, a control group was not described [10-38] and 27 added a comparison group [40-67]. The overall results provided data from 8,128 hospitalized patients with COVID-19: 4,021 TCZ-treated, in addition to standard of care (SOC) (including 711 patients admitted to ICU), and 4,107 only receiving SOC (including 694 patients admitted to ICU). SOC was basically antiviral therapy (remdesivir, lopinavir/ritonavir), antimalarials (hydroxychloroquine), or azythromicin. Characteristics of patients in the TCZ-group vs. the control group are detailed in Tables 2-7. Of the TCZ group, 2,645 (65.8%) were men with a mean age of 61.8 (SD 6.1) and median age 62.6 [range 59-65], according to the data provided. TCZ was given as a single dose in 2,030/2,952 patients (68.8%), and in 922 (31.2%), as two or more doses.

**Table 2.** Non-controlled studies. General data.

| Author | Country | Type of study | NOS | N TCZ/Control | Study period | TCZ criteria |
| --- | --- | --- | --- | --- | --- | --- |
| Xu et al.[11] | CHI | Retrospective cohort | 6 | 21/0 | FEB 5-FEB 14 | ≥30bpm or SpO2≤93% or PaO2/FiO2≤300mmHg or MV or ICU |
| Luo et al.[12] | CHI | Retrospective cohort | 6 | 15/0 | JAN 27-MAR 5 | High IL-6 and/or CRP |
| Antony et al.[13] | US | Prospective cohort | 6 | 80/0 | FEB 1-MAR 31 | O2 >3L+ PSI score ≤130 |
| Quartuccio et al.[14] | ITA | Prospective cohort | 6 | 24/0 | FEB 29*-NA | High CRP and IL-6 |
| Toniati et al.[15] | ITA | Prospective cohort | 7 | 100/0 | MAR 9-MAR 20 | BCRSS≥3 |
| Alattar et al.[16] | QAT | Retrospective cohort | 7 | 25/0 | MAR* | ≥30bpm or SpO2≤93% or PaO2/FiO2≤300mmHg +ICU+ high CRP |
| Sciascia et al.[17] | ITA | Prospective cohort | 6 | 63/0 | MAR* | SpO2<93%+PaO2/FiO2<300mmHg +<br>3 out of: CRPx10; ferritin>1,000ng/ml; D-dimerx10; LDHx2 |
| Morrison et al.[18] | US | Retrospective cohort | 7 | 81/0 | MAR 1-APR 3 | Persistent fever+ PaO2/FiO2<200mmHg+<br>1 out of:ferritin>1,000µg/l,D-dimer>5mg/ml+LDH≥500U/l, IL6x5 |
| Morena et al.[19] | ITA | Prospective cohort | 7 | 51/0 | MAR 10-MAR 23 | ≥30bpm+SpO2<93%+PaO2/FiO2<250mmHg+IL-6>40pg/ml |
| Marfella et al.[20] | ITA | Retrospective cohort | 6 | 78/0 | MAR 1-APR 10* | Moderate-to-severe respiratory illness |
| Sánchez-Montalvá et al. [21] | SPA | Retrospective cohort | 8 | 82/0 | MAR 13 -MAR 18 | PaO2/FiO2<300mmHg+ IL6>40pg/ml or D-dimer>1,500ng/ml |
| Jordan et al.[22] | US | Prospective cohort | 7 | 27/0 | MAR 13-APR 1 | SpO2<90% on 4l O2 + 2 out of: IL-6>10 pg/ml; CRP>35mg/l; Ferritin>600ng/ml; D-dimer>1mcg/l; Neutrophil-Lymphocyte Ratio>4; LDH>200U/l |
| Nasir et al.[23] | PAK | Retrospective cohort | 6 | 30/0 | FEB 26-MAY 15 | PaO2/FiO2<300 +CRP≥100mg/l or ferritin≥900ng/ml |
| Borku et al.[24] | TUR | Prospective cohort | 7 | 12/0 | MAR 18-APR 8 | increased acute phase reactants, persistent fever, decreased SpO2 and deepened respiratory distress progress |
| Issa et al.[25] | FR | Retrospective cohort | 6 | 10/0 | MAR 15-APR 30 | Persistent fever or PaO2/FiO2<300 + 2 out of: fibrinogen>8g/l, ferritin>1,000ng/ml, D-dimer>3,000ng/ml, CRP>150mg/l |
| Campins et al.[26] | SPA | Prospective cohort | 6 | 58/0 | MAR 21-APR 27 | NA |
| Sanz et al.[27] | SPA | Prospective cohort | 7 | 72/0 | MAR 15-APR 8* | 1 out of: PaO2/FiO2<300, SpO2<92, tachypnea or high ferritin levels |
| Knorr et al.[28] | US | Retrospective cohort | 7 | 66/0 | MAR 25-APR 24 | D-dimer > 1.5 mg/l |
| Lohse et al.[29] | FR | Retrospective cohort | 7 | 34/0 | APR 1-MAY 11 | ≥5l/min O2 + ≥2 out of: high ferritin/CRP/D-dimer/LDH and lymphopenia |
| Conrozier et al.[30] | FR | Retrospective cohort | 7 | 40/0 | APR 1-MAY 11 | ≥5l/min O2 + ≥2 out of: high ferritin/CRP/D-dimer/LDH and lymphopenia |
| Petrak et al.[31] | US | Retrospective cohort | 7 | 145/0 | MAR 13-APR 16 | NA |
| Górgolas et al.[32] | SPA | Retrospective cohort | 7 | 186/0 | MAR 8-APR 19 | SpO2<93% on FiO2 >24% |
| Jiménez et al.[33] | SPA | Retrospective cohort | 6 | 25/0 | MAR 26-APR 17 | ≥30bpm+SpO2≤93%+PaO2/FiO2≤300mmHg+cytokine storm |
| Rimland et al.[34] | US | Retrospective cohort | 6 | 11/0 | MAR 21-APR 25 | NA |
| Sinha et al.[35] | US | Prospective cohort | 7 | 255/0 | MAR 17-APR 30 | FiO2>45% + CRP>100mg/l, LDH>450 U/l or ferritin>700 ng/ml |
| Patel A et al.[36] | IND | Retrospective cohort | 6 | 20/0 | MAR 21-MAY 20 | SpO2<94% or CRPx10 or D-dimer>2,500ng/ml |
| Hashimoto et al. [37] | JAP | Retrospective cohort | 6 | 13/0 | NA-MAY 31 | CRP>5mg/dl or ferritin>1000ng/ml |
| Formina et al. [38] | RUS | Retrospective cohort | 6 | 89/0 | NA | NA |
| Rubio-Rivas et al. [39] | SPA | Retrospective cohort | 7 | 186/0 | MAR 17-APR 7 | PaO2/FiO2<300mmHg or SpO2/FiO2<315 + 2 out of: ferritin>1,000 ng/ml, CRP>100 mg/l, IL-6>70 ng/l, D-dimer>1,000 mcg/l, LDH>400 U/l |

\*Estimate. ALT: alanine aminotransferase. BCRSS: Brescia COVID-19 Respiratory Severity Scale. CRP: C-reactive protein. HFNC: high flow nasal cannula. ICU: intensive care unit. IL-6:interleukin 6. LDH: lactate deshydrogenase. MV: mechanical ventilation. NA: not applicable/available. NOS: Newcastle-Ottawa scale. PSI: pneumonia severity index. TCZ: tocilizumab

**Table 3.** Controlled studies. General data.

| Author | Country | Type of study | NOS | N TCZ/Control | Study period | TCZ criteria |
| --- | --- | --- | --- | --- | --- | --- |
| Rodríguez-Baño et al. [40] | SPA | Retrospective cohort | 8 | 88/344<br>151/344 | FEB 2-MAR 31 | ≥38°C + increase in oxygen support to maintain SpO<sub>2</sub>>92%+ 1 out of:<br>ferritin>2,000ng/ml, D-dimer>1,500mcg/ml, IL-6>50pg/ml |
| Quartuccio et al.[41] | ITA | Retrospective cohort | 8 | 42/69 | FEB 29-APR 6 | High CRP and IL-6 |
| Martínez-Sanz et al.[42] | SPA | Retrospective cohort | 7 | 260/969 | JAN 31-APR 23 | NA |
| Moreno-García et al.[43] | SPA | Retrospective cohort | 8 | 77/94 | FEB 19-APR 16 | CRP≥8mg/dl, ferritin≥800ng/ml or lymphocyte count<800cells/mm<sup>3</sup> |
| Guaraldi et al.[44] | ITA | Retrospective cohort | 8 | 179/365 | FEB 21-APR 30 | ≥30bpm+SpO<sub>2</sub><93%+PaO<sub>2</sub>/FiO<sub>2</sub><300mmHg |
| Campochiaro et al.[45] | ITA | Retrospective cohort | 8 | 32/33 | MAR 13-MAR 19 | SpO<sub>2</sub>≤92% + PaO<sub>2</sub>/FiO<sub>2</sub> ≤300 mmHg +<br>LDH>220 U/l + CRP≥100mg/l or ferritin ≥900 ng/ml |
| Mikulska et al.[46] | ITA | Retrospective cohort | 7 | 85/66 | MAR 11-MAR 24 | ≥30bpm+SpO<sub>2</sub>≤93%+PaO<sub>2</sub>/FiO<sub>2</sub>≤300mmHg+cytokine storm |
| Colaneri et al.[47] | ITA | Retrospective cohort | 8 | 21/91 | MAR 14-MAR 27 | CRP>5 mg/dl+Procalcitonin<0.5 ng/mL+PaO<sub>2</sub>/FiO<sub>2</sub><300mmHg+ALT<500 U/L. |
| Price et al.[48] | US | Retrospective cohort | 8 | 153/86 | MAR 10-MAR 31 | O<sub>2</sub> ≥ 3 l to maintain SpO<sub>2</sub>>93% or MV |
| Maeda et al.[49] | US | Retrospective cohort | 6 | 23/201 | MAR 13-MAR 31 | NA |
| Capra et al.[50] | ITA | Retrospective cohort | 8 | 62/23 | MAR 13-APR 2 | 1 out of:≥30bpm+SpO<sub>2</sub>≤93%+PaO<sub>2</sub>/FiO<sub>2</sub>≤300mmHg |
| Rossotti et al.[51] | ITA | Retrospective cohort | 8 | 74/148 | MAR 13-APR 3 | ≥30bpm+SpO<sub>2</sub>≤93%+PaO<sub>2</sub>/FiO<sub>2</sub>≤300mmHg or ICU +<br>CRP>1 mg/dl or IL-6>40 pg/ml or D-dimer >1.5 mcg/ml or ferritin >500 ng/ml |
| Eimer et al.[52] | SWE | Retrospective cohort | 8 | 29/58 | MAR 11-APR 15 | SpO<sub>2</sub><94 on 5l O<sub>2</sub> + 1 out of: CRP>100mg/L, LDH>8 µkat/L, IL-6>40 ng/L, D-Dimer>2mg/L, troponin T>15 ng/L,ferritin>500 µg/L |
| Kimmig et al.[53] | US | Retrospective cohort | 7 | 48/63 | MAR 1-APR 27 | Progressive clinical deterioration + inflammation markers |
| Somers et al.[54] | US | Prospective cohort | 7 | 78/76 | MAR 9-APR 20 | MV |
| Tsai et al.[55] | US | Retrospective cohort | 7 | 66/66 | MAR 1-MAY 5 | SpO<sub>2</sub>≤94%+ferritin>300 mcg/ml |
| Kewan et al.[56] | US | Retrospective cohort | 7 | 28/23 | MAR 13-APR 19 | CRP≥3g/dl or ferritin>400ng/ml |
| Patel K et al.[57] | US | Retrospective cohort | 7 | 42/41 | MAR 16-APR 17 | NA |
| Wadud et al.[58] | US | Retrospective cohort | 6 | 44/50 | MAR 15-APR 20 | NA |
| Ramaswamy et al.[59] | US | Retrospective cohort | 7 | 21/65 | MAR 16-APR 22 | SpO<sub>2</sub>≤88%+CRP≥7mg/dl |
| Rojas-Marte et al.[60] | US | Retrospective cohort | 8 | 96/97 | MAR 8-APR 25 | O<sub>2</sub> mask/HFNC up to 10l to maintain SpO<sub>2</sub>≥95% or MV |
| Roumier et al.[61] | FR | Retrospective cohort | 8 | 30/29 | MAR 21-APR 2 | >6l O<sub>2</sub> + high CRP |
| Rossi et al.[62] | FR | Retrospective cohort | 7 | 106/140 | MAR 23-NA | SpO<sub>2</sub>≤96%+on 6l O<sub>2</sub> |
| Potere et al.[63] | ITA | Retrospective cohort | 7 | 40/40 | MAR 28-APR 21 | Pneumonia+CRP≥20mg/dl+SpO<sub>2</sub><90% |
| Klopfenstein et al.[64] | FR | Retrospective cohort | 8 | 20/25 | APR 1-APR 13 | ≥5l/min O<sub>2</sub> + ≥2 out of: high ferritin/CRP/D-dimer/LDH and lymphopenia |
| Canziani et al.[65] | ITA | Retrospective cohort | 7 | 64/64 | FEB 23-MAY 9 | Respiratory worsening + High CRP, ferritin, CK, ALT, D-dimer or lymphopenia |
| Ip et al.[66] | US | Retrospective cohort | 7 | 134/413 | APR 22-MAY 5 | NA |
| Carvalho et al.[67] | BRA | Prospective cohort | 7 | 29/24 | MAR 21-MAY 31 | ICU + fever + High CRP≥5mg/dl |

ALT: alanine aminotransferase. BCRSS: Brescia COVID-19 Respiratory Severity Scale. CRP: C-reactive protein. HFNC: high flow nasal cannula. ICU: intensive care unit. IL-6:interleukin 6. LDH: lactate deshydrogenase. MV: mechanical ventilation. NA: not applicable/available. NOS: Newcastle-Ottawa scale. PSI: pneumonia severity index. TCZ: tocilizumab

**Table 4.**
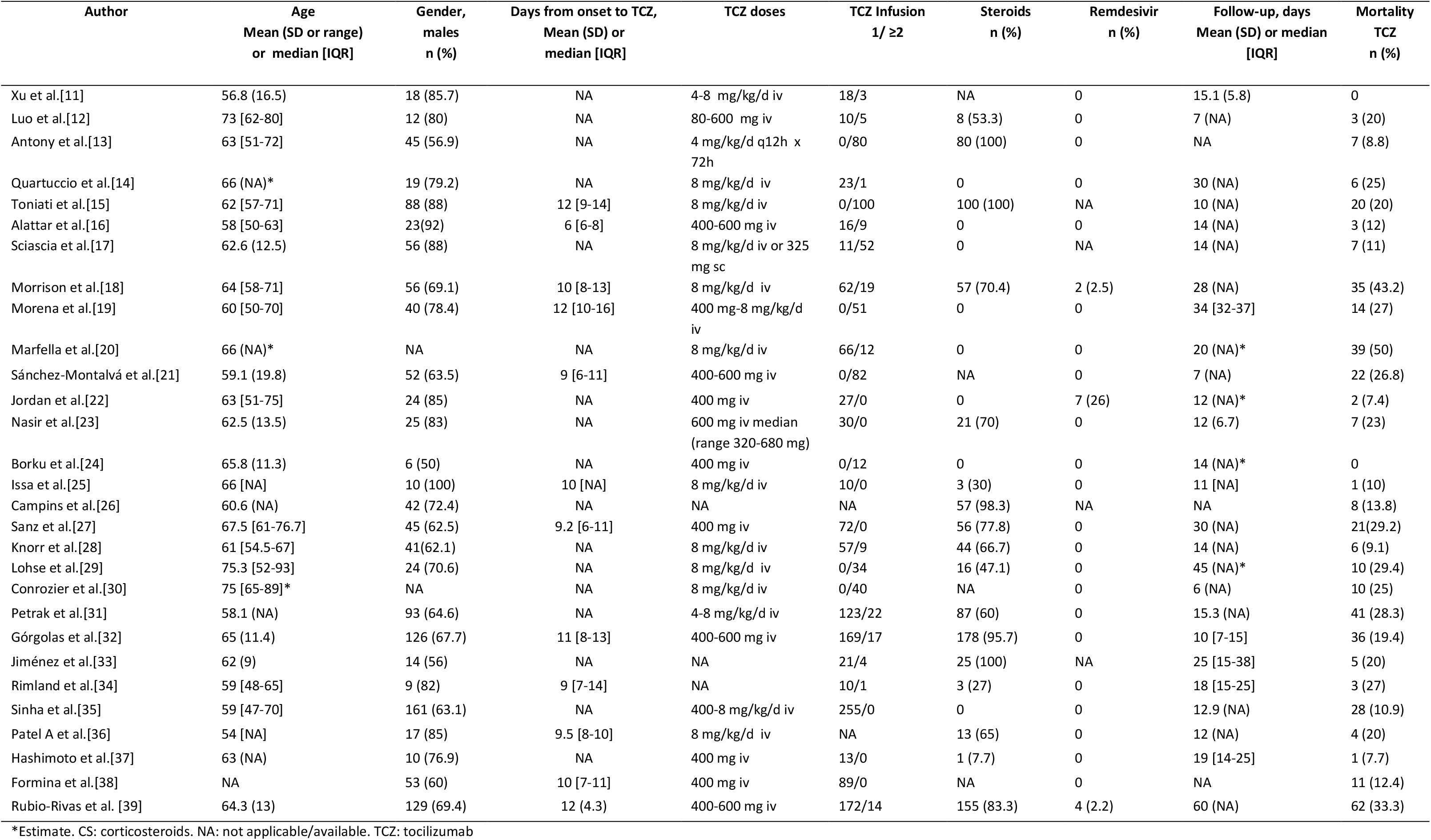
Non-controlled studies. Drugs and mortality.

| Author | Age<br>Mean (SD or range)<br>or median [IQR] | Gender,<br>males<br>n (%) | Days from onset to TCZ,<br>Mean (SD) or<br>median [IQR] | TCZ doses | TCZ Infusion<br>1/ ≥2 | Steroids<br>n (%) | Remdesivir<br>n (%) | Follow-up, days<br>Mean (SD) or median<br>[IQR] | Mortality<br>TCZ<br>n (%) |
| --- | --- | --- | --- | --- | --- | --- | --- | --- | --- |
| Xu et al.[11] | 56.8 (16.5) | 18 (85.7) | NA | 4-8 mg/kg/d iv | 18/3 | NA | 0 | 15.1 (5.8) | 0 |
| Luo et al.[12] | 73 [62-80] | 12 (80) | NA | 80-600 mg iv | 10/5 | 8 (53.3) | 0 | 7 (NA) | 3 (20) |
| Antony et al.[13] | 63 [51-72] | 45 (56.9) | NA | 4 mg/kg/d q12h x<br>72h | 0/80 | 80 (100) | 0 | NA | 7 (8.8) |
| Quartuccio et al.[14] | 66 (NA)* | 19 (79.2) | NA | 8 mg/kg/d iv | 23/1 | 0 | 0 | 30 (NA) | 6 (25) |
| Toniati et al.[15] | 62 [57-71] | 88 (88) | 12 [9-14] | 8 mg/kg/d iv | 0/100 | 100 (100) | NA | 10 (NA) | 20 (20) |
| Alattar et al.[16] | 58 [50-63] | 23(92) | 6 [6-8] | 400-600 mg iv | 16/9 | 0 | 0 | 14 (NA) | 3 (12) |
| Sciascia et al.[17] | 62.6 (12.5) | 56 (88) | NA | 8 mg/kg/d iv or 325<br>mg sc | 11/52 | 0 | NA | 14 (NA) | 7 (11) |
| Morrison et al.[18] | 64 [58-71] | 56 (69.1) | 10 [8-13] | 8 mg/kg/d iv | 62/19 | 57 (70.4) | 2 (2.5) | 28 (NA) | 35 (43.2) |
| Morena et al.[19] | 60 [50-70] | 40 (78.4) | 12 [10-16] | 400 mg-8 mg/kg/d<br>iv | 0/51 | 0 | 0 | 34 [32-37] | 14 (27) |
| Marfella et al.[20] | 66 (NA)* | NA | NA | 8 mg/kg/d iv | 66/12 | 0 | 0 | 20 (NA)* | 39 (50) |
| Sánchez-Montalvá et al.[21] | 59.1 (19.8) | 52 (63.5) | 9 [6-11] | 400-600 mg iv | 0/82 | NA | 0 | 7 (NA) | 22 (26.8) |
| Jordan et al.[22] | 63 [51-75] | 24 (85) | NA | 400 mg iv | 27/0 | 0 | 7 (26) | 12 (NA)* | 2 (7.4) |
| Nasir et al.[23] | 62.5 (13.5) | 25 (83) | NA | 600 mg iv median<br>(range 320-680 mg) | 30/0 | 21 (70) | 0 | 12 (6.7) | 7 (23) |
| Borku et al.[24] | 65.8 (11.3) | 6 (50) | NA | 400 mg iv | 0/12 | 0 | 0 | 14 (NA)* | 0 |
| Issa et al.[25] | 66 [NA] | 10 (100) | 10 [NA] | 8 mg/kg/d iv | 10/0 | 3 (30) | 0 | 11 [NA] | 1 (10) |
| Campins et al.[26] | 60.6 (NA) | 42 (72.4) | NA | NA | NA | 57 (98.3) | NA | NA | 8 (13.8) |
| Sanz et al.[27] | 67.5 [61-76.7] | 45 (62.5) | 9.2 [6-11] | 400 mg iv | 72/0 | 56 (77.8) | 0 | 30 (NA) | 21(29.2) |
| Knorr et al.[28] | 61 [54.5-67] | 41(62.1) | NA | 8 mg/kg/d iv | 57/9 | 44 (66.7) | 0 | 14 (NA) | 6 (9.1) |
| Lohse et al.[29] | 75.3 [52-93] | 24 (70.6) | NA | 8 mg/kg/d iv | 0/34 | 16 (47.1) | 0 | 45 (NA)* | 10 (29.4) |
| Conrozier et al.[30] | 75 [65-89]* | NA | NA | 8 mg/kg/d iv | 0/40 | NA | 0 | 6 (NA) | 10 (25) |
| Petrak et al.[31] | 58.1 (NA) | 93 (64.6) | NA | 4-8 mg/kg/d iv | 123/22 | 87 (60) | 0 | 15.3 (NA) | 41 (28.3) |
| Górgolas et al.[32] | 65 (11.4) | 126 (67.7) | 11 [8-13] | 400-600 mg iv | 169/17 | 178 (95.7) | 0 | 10 [7-15] | 36 (19.4) |
| Jiménez et al.[33] | 62 (9) | 14 (56) | NA | NA | 21/4 | 25 (100) | NA | 25 [15-38] | 5 (20) |
| Rimland et al.[34] | 59 [48-65] | 9 (82) | 9 [7-14] | NA | 10/1 | 3 (27) | 0 | 18 [15-25] | 3 (27) |
| Sinha et al.[35] | 59 [47-70] | 161 (63.1) | NA | 400-8 mg/kg/d iv | 255/0 | 0 | 0 | 12.9 (NA) | 28 (10.9) |
| Patel A et al.[36] | 54 [NA] | 17 (85) | 9.5 [8-10] | 8 mg/kg/d iv | NA | 13 (65) | 0 | 12 (NA) | 4 (20) |
| Hashimoto et al.[37] | 63 (NA) | 10 (76.9) | NA | 400 mg iv | 13/0 | 1 (7.7) | 0 | 19 [14-25] | 1 (7.7) |
| Formina et al.[38] | NA | 53 (60) | 10 [7-11] | 400 mg iv | 89/0 | NA | 0 | NA | 11 (12.4) |
| Rubio-Rivas et al. [39] | 64.3 (13) | 129 (69.4) | 12 (4.3) | 400-600 mg iv | 172/14 | 155 (83.3) | 4 (2.2) | 60 (NA) | 62 (33.3) |

\*Estimate. CS: corticosteroids. NA: not applicable/available. TCZ: tocilizumab

**Table 5.**
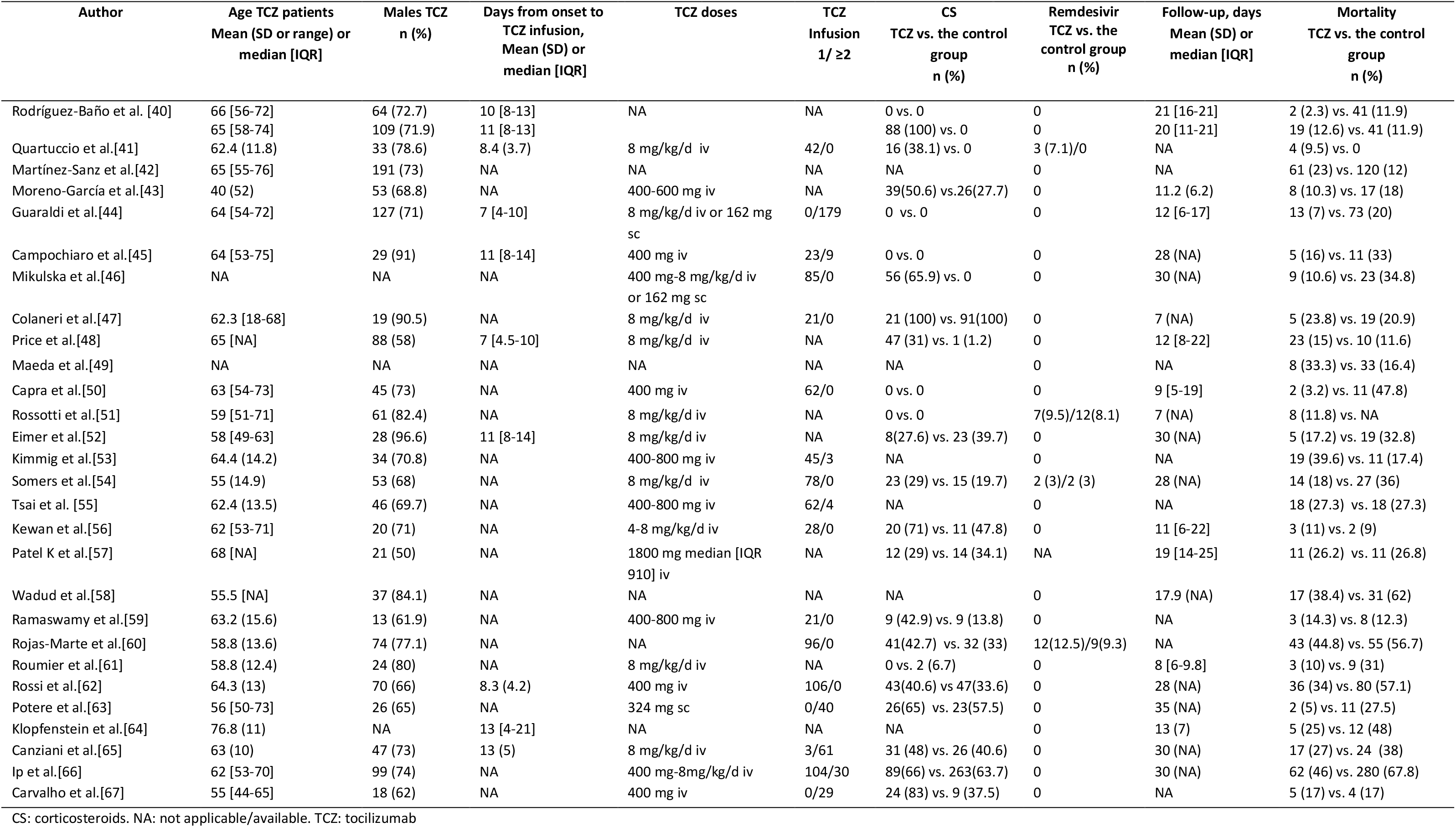
Controlled studies. Drugs and mortality.

**Table 6.** Non-controlled studies. Length of hospital stay, Mortality ICU vs. ward and secondary infections.

| Author | Length of stay<br>Days, mean (SD) or median [IQR] | ICU admission TCZ<br>n/N (%) | TCZ prescribed at ICU<br>mortality n/N (%) | TCZ prescribed at the ward<br>mortality n/N (%) | Secondary Infections<br>n/N (%) |
| --- | --- | --- | --- | --- | --- |
| Xu et al. [11] | 15.1 (5.8) | 2/20 (10) | NA | 0/20 (0) | Overall 0/20 (0) |
| Luo et al. [12] | NA | NA | 3/7 (42.9) | 0/8 (0) | NA |
| Antony et al. [13] | NA (range 5-10) | 9/80 (11.3) | NA | 7/80 (8.7) | Overall 12/80 (15)<br>Bacterial 12/80 (15) |
| Quartuccio et al. [14] | NA | NA | NA | NA | NA |
| Toniati et al. [15] | NA | 3/57 (5.3) | 10/43 (24) | 10/57 (17.5) | Overall 2/100 (2)<br>Bacterial 2/100 (2) |
| Alattar et al. [16] | NA | NA | 3/25 (12) | NA | Overall 8/25 (32)<br>Fungal 8/25 (32) |
| Sciascia et al. [17] | NA | NA | 1/5 (20) | 6/58 (10.3) | NA |
| Morrison et al. [18] | 18 [13-29] | NA | 32/70 (45.7) | 3/11 (27.3) | Overall 20/81 (24.7)<br>Bacterial 17/81 (21)<br>Fungal 3/81 (3.7) |
| Morena et al. [19] | NA | NA | 5/6 (83) | 9/45 (20) | Overall 14/51 (27)<br>Bacterial 14/51 (27) |
| Marfella et al. [20] | NA | NA | NA | NA | NA |
| Sánchez-Montalvá et al. [21] | NA | 14/82 (17.1) | NA | 22/82 (26.8) | Overall 0/82 (0) |
| Jordan et al. [22] | NA | 1/27 (3.7) | 2/21 (9.5) | 0/6 (0) | Overall 2/27 (7.4)<br>Bacterial 2/27 (7.4) |
| Nasir et al. [23] | 12 (6.7) | 0/20 (0) | NA | NA | Overall 16/30 (54)<br>Bacterial 9/30 (30)<br>Fungal 7/30 (24) |
| Borku et al. [24] | NA | 2/12 (16.7) | NA | 0/12 (0) | NA |
| Issa et al. [25] | NA | 0 | 1/10 (10) | NA | Overall 2/10 (20)<br>Bacterial 1/10 (10)<br>Fungal 1/10 (10) |
| Campins et al. [26] | NA | 19/58 (32.4) | NA | 8/58 (13.8) | NA |
| Sanz et al. [27] | 16.4 [11-20] | 43/72 (59.7) | NA | 21/72 (29.2) | NA |
| Knorr et al. [28] | 16.4 [8.8-23] | 16/48 (33.3) | 1/18 (5.6) | 3/48 (6.25) | NA |
| Lohse et al. [29] | NA | NA | NA | 10/34 (29.4) | NA |
| Conrozier et al. [30] | NA | NA | NA | NA | NA |
| Petrak et al. [31] | 15.3 (NA) | NA | 35/81 (43.2) | 6/64 (9.4) | Overall 0/145 (0) |
| Górgolas et al. [32] | NA | 19/186 (10.2) | NA | 36/186 (19.4) | Overall 13/186 (6.3)<br>Bacterial 6/186 (3.2)<br>Fungal 9/186 (4.8) |
| Jiménez et al. [33] | 25 [15-38] | NA | NA | NA | Overall 8/25 (32)<br>Bacterial 8/25 (32) |
| Rimland et al. [34] | NA | 5/7 (71.4) | NA | NA | Overall 2/11 (18.2)<br>Bacterial 2/11 (18.2) |
| Sinha et al. [35] | 12.9 (NA) | NA | NA | NA | Overall 34/255 (13.3)<br>Bacterial 34/255 (13.3) |
| Patel A et al. [36] | NA | 2/20 (10) | NA | 4/20 (20) | Overall 1/20 (5)<br>Bacterial 1/20 (5) |
| Hashimoto et al. [37] | 17 (range 12-27) | 4/11 (36.4) | 0/2 (0) | 1/11 (9.1) | NA |
| Formina et al. [38] | NA | NA | 10/17 (58.8) | 1/72 (1.4) | NA |
| Rubio-Rivas et al. [39] | NA | NA | NA | NA | NA |

ICU: intensive care unit. NA: Not available/applicable. TCZ: tocilizumab

**Table 7.** Controlled studies. Length of hospital stay, Mortality ICU vs. ward and secondary infections.

|  |  |  |  |  |  |
| --- | --- | --- | --- | --- | --- |
| Author | Length of stay TCZ vs. Control, days, mean (SD) or median [IQR] | ICU admission TCZ vs. Control, n/N (%) | TCZ prescribed at ICU mortality TCZ vs. Control, n/N (%) | TCZ prescribed at the ward mortality TCZ vs. Control, n/N (%) | Secondary Infections TCZ vs. control group n/N (%) |
| Rodriguez-Baño et al.[40] | NA | NA | NA | NA | Overall 11/88 (12.5) vs. 36/339 (10.3)<br>Bacterial 11/88 (12.5) vs. 36/339 (10.3)<br>Overall 18/150 (12) vs. 36/339 (10.3)<br>Bacterial 18/150 (12) vs. 36/339 (10.3) |
| Quartuccio et al.[41] | NA | 0/15 (0) vs. 0/69 (0) | 3/27 (11.1) vs. 0/0 (0) | 1/15 (6.7) vs. 0/69 (0) | Overall 18/27 (16.2) vs. 0 (0)<br>Bacterial 18/27 (16.2) vs. 0 (0) |
| Martínez-Sanz et al.[42] | 19 [NA] vs. 10 [NA]* | 50/260 (19) vs. 32/969 (3) | NA | 61/260 (23) vs. 120/969 (12) | NA |
| Moreno-García et al.[43] | NA | 8/77 (10.3) vs. 26/94 (27.6) | NA | 8/77 (10.3) vs. 17/94 (18) | NA |
| Guaraldi et al.[44] | NA | NA | NA | NA | Overall 24/132 (14.4) vs. 14/222 (6)<br>Bacterial 12/132 (9.1) vs.11/222 (4.6)<br>Fungal 7/132 (5.3) vs. 3/222 (1.4) |
| Campochiaro et al.[45] | 13.5[10-16.7] vs. 14 [12-15.5] | 4/32 (13) vs. 2/33 (6) | NA | 5/32 (16) vs. 11/33 (33) | Overall 5/32 (16.1) vs. 4/33 (12)<br>Bacterial 4/32 (13) vs. 4/33 (12)<br>Fungal 1/32 (3.1) vs. 0/33 (0) |
| Mikulska et al.[46] | NA | NA | NA | NA | NA |
| Colaneri et al.[47] | 2[6] vs. 14 [4] | 3/21 (14.3) vs. 12/91 (13.2) | NA | 5/21 (23.8) vs. 19/91 (20.9) | Overall 0/21 (0) vs. 0/91 (0) |
| Price et al.[48] | 12[8-22] vs. NA | NA | NA | NA | Overall 4/153 (2.6) vs.NA<br>Bacterial 4/153 (2.6) vs.NA |
| Maeda et al.[49] | NA | NA | NA | NA | Overall 3/23 (13) vs. 2/201 (1.1)<br>Fungal 3/23 (13) vs. 2/201 (1.1) |
| Capra et al.[50] | 12.5 [4-18] vs. 8 [7-15] | NA | NA | NA | Overall 0/62 (0) vs. NA |
| Rossotti et al.[51] | NA | NA | NA | NA | Overall 24/74 (32.4) vs. NA |
| Eimer et al.[52] | 20.5 [16.5-30] vs. 30[21.5-30] | NA | 5/29 (17.2) vs. 19/58 (32.8) | NA | Overall 5/29 (17.2) vs. 14/58 (24.1)<br>Bacterial 5/29 (17.2) vs. 14/58 (24.1) |
| Kimmig et al.[53] | NA | NA | 19/48 (39.6) vs. 11/63 (17.4) | NA | Overall 27/48 (56.3) vs. 18/63 (28.6)<br>Bacterial 24/48 (50) vs. 18/63 (28.6)<br>Fungal 3/48 (6.3) vs. 0/63 (0) |
| Somers et al.[54] | 20.4 [13.8-35.8] vs. 22.9[16.3-28.5] | NA | 14/78 (18) vs. 27/76 (36) | NA | Overall 41/78 (52.5) vs. 22/76 (28.9)<br>Bacterial 38/78 (48.7) vs. 21/76 (27.6)<br>Fungal 3/78 (3.8) vs. 1/76 (1.3) |
| Tsai et al.[55] | NA | NA | NA | NA | NA |
| Kewan et al.[56] | 11[6-22.3] vs. 7 [5-13.5] | NA | NA | NA | Overall 5/28 (18) vs. 5/51 (22) |
| Patel K et al.[57] | NA | NA | NA | NA | NA |
| Wadud et al.[58] | NA | NA | NA | NA | NA |
| Ramaswamy et al.[59] | NA | NA | NA | NA | NA |
| Rojas-Marte et al.[60] | 14.5 (8.8) vs. 16.5 (10.8) | NA | 41/61 (67.2) vs. 45/60 (75)* | 2/33(6.1) vs. 9/34 (26.5)* | Overall 16/61 (16.7) vs. 26/60 (26.8)<br>Bacterial 12/61 (12.5) vs. 23/60 (23.7)<br>Fungal 4/61 (4.2) vs.3/60 (3.1) |
| Roumier et al.[61] | NA | NA | NA | NA | NA |
| Rossi et al.[62] | NA | NA | NA | NA | NA |
| Potere et al.[63] | NA | NA | NA | NA | Overall 1/40 (2.5) vs. 3/40 (7.5)<br>Bacterial 1/40 (2.5) vs. 3/40 (7.5) |
| Klopfenstein et al.[64] | 13 [4-32] vs. 17 [5-41] | 0/20 (0) vs. 11/25 (44) | NA | 5/20 (25) vs. 12/25 (48) | NA |
| Canziani et al.[65] | NA | NA | NA | NA | Overall 20/64 (31) vs. 25/64 (39) |
| Ip et al.[66] | NA | NA | 62/134 (46) vs. 231/413 (56) | NA | Overall 30/134 (22) vs. 69/413 (17)<br>Bacterial 30/134 (22) vs. 69/413 (17) |
| Carvalho et al.[67] | NA | NA | 5/29 (17) vs. 4/24 (17) | NA | Overall 16/29 (55) vs. 4/24 (16)<br>Bacterial 10/29 (34) vs. 3/24 (12)<br>Fungal 6/29 (21) vs. 1/24 (4) |

\*Estimate. ICU: intensive care unit. NA: Not available/applicable. TCZ: tocilizumab.

Concomitantly to TCZ use, additional treatment with steroids was given in 1,560/3,073 patients in the TCZ-group (50.8%) vs. 592/2,733 (21.7%) in the control group (p<0.001). Comparing both groups, remdesivir was used in 37/3,511 (1.05%) vs. 23/3,945 (0.58%) (p=0.023) patients. Finally, the administration of TCZ has prescribed a median of 10 days [range 9-11] after the onset of COVID-19 symptoms, in those studies in which this data was provided. The median follow-up of the overall cohort was 10.3 days [range 12-19].

### Mortality

After applying the random-effects model, hospital-wide (including ICUs) pooled mortality of patients with COVID-19 treated with TCZ was 19.2% (95%CI 16.4-22.5) (I^2^=83.6% Q=305.3 tau^2^=0.23 p<0.001). Egger’s method A=-3.354 p<0.001 (Figure 3). In the control group, overall mortality was 27.4% (95%CI 21.1-35.6%) (I^2^=95.9% Q=569.1 tau^2^=0.38 p<0.001). These differences between the TCZ-group and the control group achieved statistical significance (p<0.001). The RR of mortality in the TCZ-group was 0.73 (95%CI 0.57-0.93; p=0.010) (I^2^=77.7% Q=107.4 tau^2^=0.24 p<0.001). Egger’s method A=- 0.712 p=0.380 (Figure 4). The number needed to treat (NNT) to avoid one death was 20.

**Figure 3.**
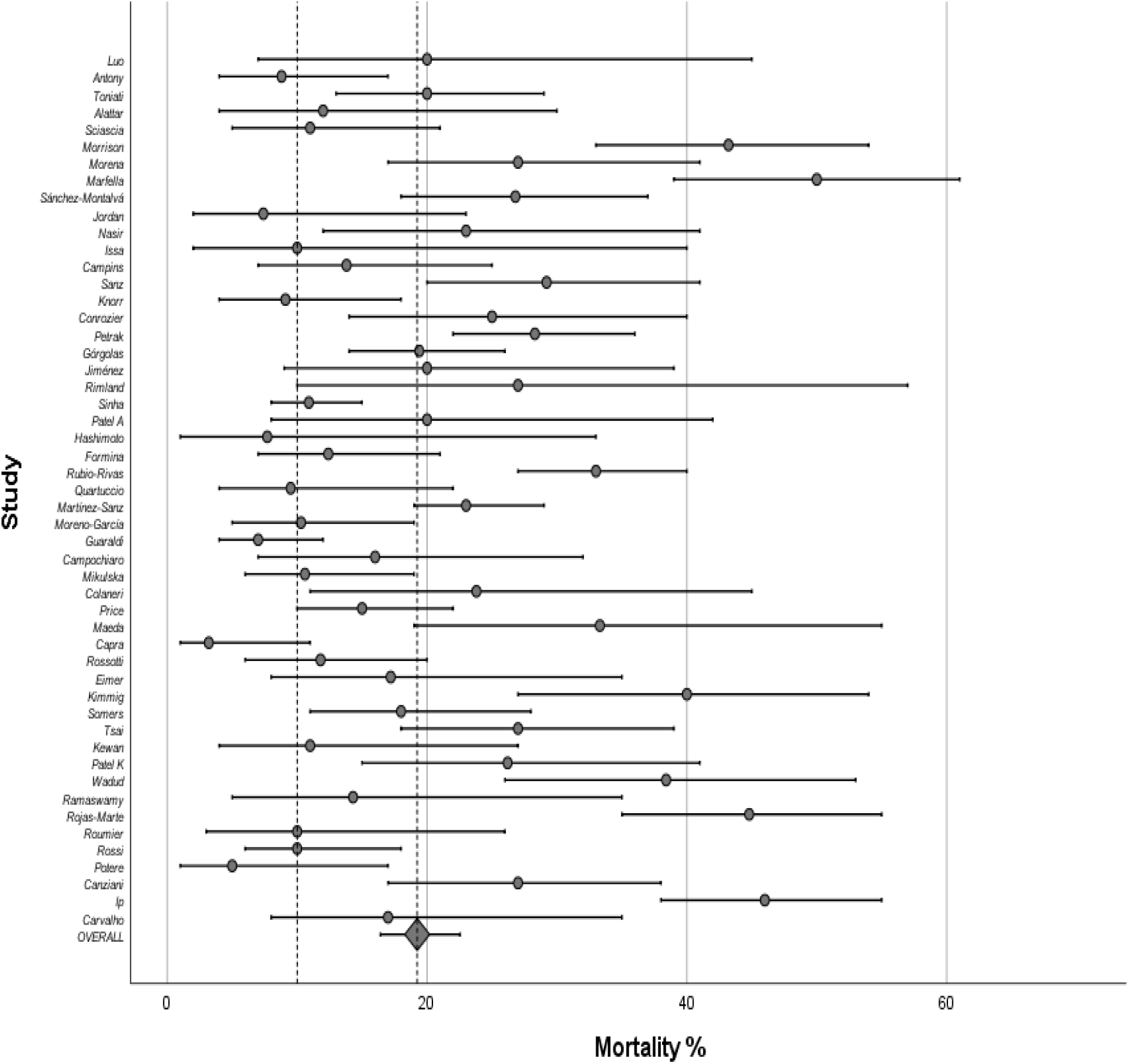
Mortality (percentage) in COVID-19 patients receiving TCZ. Forest plot.

**Figure 4.**
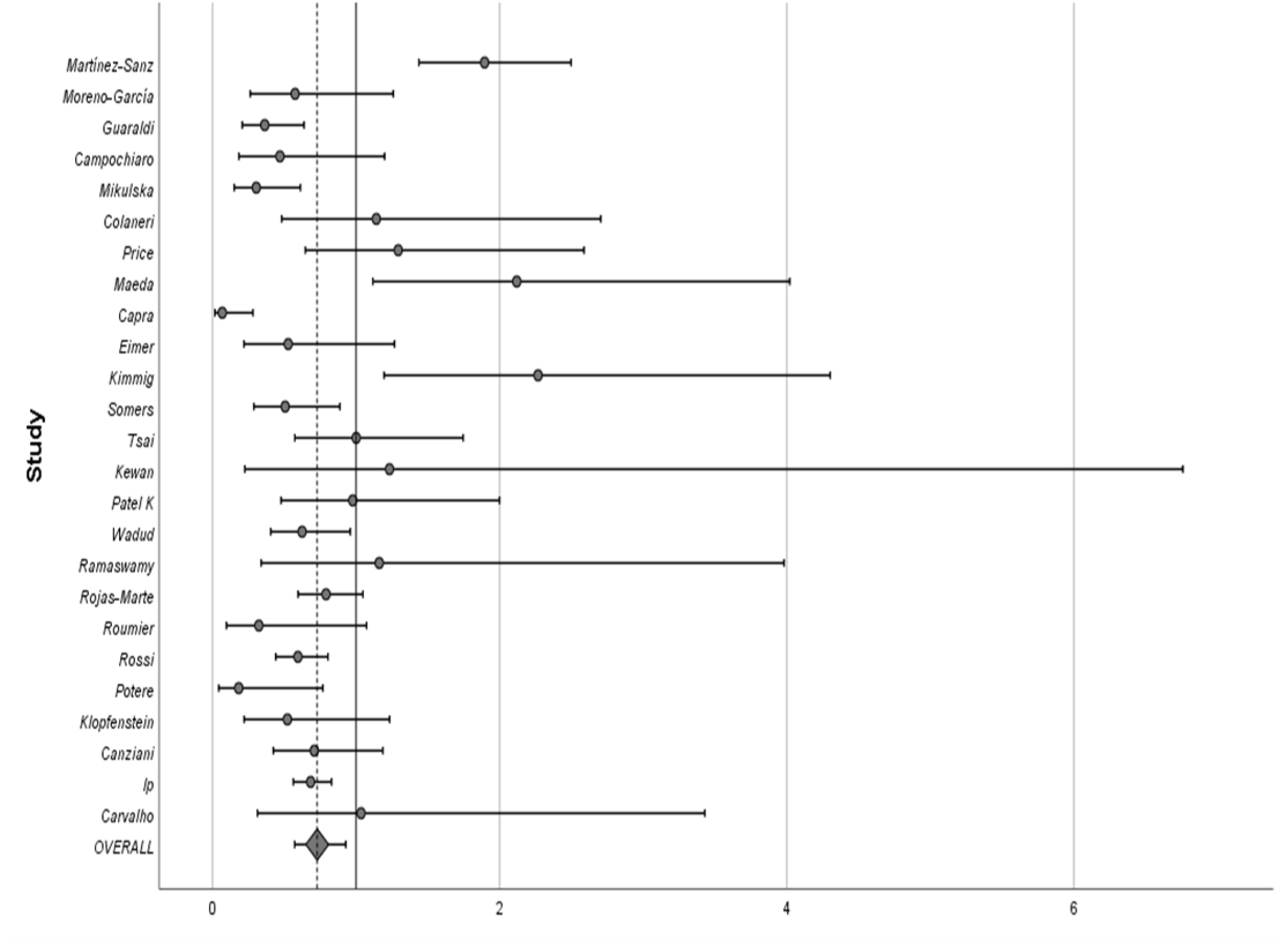
Mortality (RR) in the TCZ-group vs. the control group. Forest plot.

When the analysis was restricted to high-quality controlled observational studies, excluding those non-controlled, those with <20 patients, or those showing NOS <7, the pooled mortality in the TCZ-group was 17.7% (95%CI 13.9-22.6) (I^2^=85% Q=168 tau^2^=0.295 p<0.001). Egger’s method A=-3.727 p<0.001. The RR of death in the TCZ-group vs. the control group was 0.58 (95%CI 0.40-0.85) (I^2^=79.7% Q=108.3 tau^2^=0.60 p=0.004). Egger’s method A=-1.511 p=0.153.

#### Overall Mortality in hospital wards

The pooled mortality of COVID-19 patients receiving TCZ in conventional wards was 17% (95%CI 13.9-20.8) (I^2^=58.5% Q=50.6 tau2=0.108 p<0.001). Egger’s method A=-2.082 p<0.001. In contrast, mortality in the control group after being admitted for COVID-19 in conventional wards was 23.8% (95%CI 14.8-38.4) (I^2^=90.3% Q=51.8 tau^2^=0.314 p<0.001). Egger’s method A=5.433 p=0.044. These differences did not achieve statistical significance (p=0.261). The RR of mortality in hospital wards in the TCZ-group vs. the control group was 0.64 (95%CI 0.26-1.54; p=0.314) (I^2^=80.8% Q=26 tau^2^=0.91 p=0.314). Egger’s method A=-3.576 p=0.005. When analysis was restricted to high-quality controlled studies, the pooled mortality in hospital wards in the TCZ-group was 13.3% (95%CI 8.7-20.4) (I^2^=76.2% Q=25 tau^2^=0.235 p<0.001). Egger’s method A=-2.653 p=0.074.

#### Overall Mortality in ICUs

The pooled mortality of COVID-19 patients receiving TCZ once admitted to ICU was 31.7% (95%CI 24.7-40.8) (I^2^=82.6% Q=97.6 tau^2^=0.185 p<0.001). Egger’s method A=-2.995 p<0.001. When compared, mortality of ICU control group patients was 39.1% (95%CI 29.1-52-4) (I^2^=91.6% Q=59.4 tau^2^=0.101 p<0.001). Egger’s method A=-1.766 p=0.290. These differences achieved statistical significance (p<0.001). The RR for mortality in those ICU patients receiving TCZ vs. SOC was 0.78 (95%CI 0.44-1.35; p=0.369) (I^2^=65.4% Q=14.4 tau^2^=0.29 p=0.639). Egger’s method A=0.693 p=0.727. The NNT in ICU to avoid one death was 9.

When the analysis was restricted to high-quality controlled studies, the pooled mortality in the ICU in the TCZ-group was 29.3% (95%CI 19.4-44.2) (I^2^=89.1% Q=55 tau^2^=0.235 p<0.001). Egger’s method A=-4.564 p=0.009.

#### Mortality in the TCZ-group receiving additional corticosteroids

Concomitantly to TCZ use, additional treatment with steroids was given in 1,560/3,073 patients in the TCZ-group (50.8%) vs. 592/2,733 (21.7%) in the control group (p<0.001). Since outcomes were not reported separately in most of the included studies, differences in mortality could only be compared between those studies in which steroids were not prescribed in addition of TCZ use and those in which 100% of TCZ-treated patients received concomitant therapy with steroids.

The pooled mortality of TCZ patients without corticosteroids was 10.8% (95%CI 6.3-18.5) (I^2^=91% Q=136.3 tau^2^=0.820 p<0.001). Egger’s method A=-4.438 p=0.006. ***Alternatively,*** the pooled mortality of TCZ patients with corticosteroids was 16.3% (95%CI 12.8-20.8) (I^2^=35.9% Q=6.2 tau^2^=0.046 p<0.001). Egger’s method A=-0.342 p=0.895.

#### Mortality in the TCZ-group receiving early vs. late TCZ administration

Hospital-wide (including ICUs) pooled mortality in TCZ-treated patients in whom TCZ was early administered (<10 days from symptoms onset) was 15.9% (95%CI 10.9-23) (I^2^=74.9% Q=31.9 tau^2^=0.222 p<0.001). Egger’s method A=-2.306 p=0.239. Hospital-wide (including ICUs) pooled mortality in patients with COVID-19 illness in whom TCZ was administered later (≥10 days) was 23.3% (95%CI 17.9-30.3) (I^2^=75.5% Q=36.8 tau^2^=0.115 p<0.001). Egger’s method A=-2.983 p=0.021. Differences between groups did not achieve statistical significance (p=0.252).

When the analysis was restricted to high-quality controlled studies, the pooled mortality after early TCZ was 11% (95%CI 8.4-14.3) (I^2^=42.4% Q=5.2 tau^2^=0.057 p<0.001). Egger’s method A=-2.824 p=0.372. Alternatively, the pooled mortality after late TCZ was 22.6% (95%CI 16.4-31.1) (I^2^=3.4% Q=2.1 tau^2^=0.004 p<0.001). Egger’s method A=-2.756 p=0.034.

### Risk of ICU admission

In patients with COVID-19 initially admitted to hospital wards, the pooled ICU admission rate after TCZ administration was 17.1% (95%CI 11.5-25.5) (I^2^=90% Q=149.5 tau^2^=0.51 p<0.001). Egger’s method A=-3.272 p=0.015. Conversely, the risk for ICU admission in the control group, initially admitted outside the ICUs, was 9.5% (95%CI 2.9-31.2) (I^2^=96.2% Q=78.8 tau^2^=1.37 p<0.001). Egger’s method A=-1.315 p=0.886. These differences achieved statistical significance (p<0.001). Accordingly, in the subset of patients initially admitted outside the ICUs, those receiving TCZ showed a RR of ICU admission of 1.49 (95%CI 0.30-7.34; p=0.621) (I^2^=93% Q=43 tau^2^=2.35 p=0.62). Egger’s method A=-4.026 p=0.435.

When the analysis was restricted to high-quality controlled studies, the pooled risk of ICU admission after receiving TCZ at the ward was 15.6% (95%CI 11.8-20.9) (I^2^=22.4% Q=3 tau^2^=0.022 p<0.001). Egger’s method A=-1.839 p=0.113.

### Safety

The pooled rate of reported secondary viral, bacterial or opportunistic fungal infections in those patients with COVID-19 treated with TCZ was 18.9% (95%CI 14.5-24.8) (I^2^=88.1% Q=218.8 tau^2^=0.391 p<0.001). Egger’s method A=-3.852 p=0.001. The RR of secondary infections in TCZ-treated vs. the control group was 1.47 (95%CI 0.99-2.19; p=0.058) (I^2^=66.1% Q=38.3 tau^2^=0.34 p=0.058). Egger’s method A=0.039 p=0.977

When the analysis was restricted to high-quality controlled studies, the pooled percentage of secondary infections was 20.7% (95%CI 14.6-29.2) (I^2^=88.2% Q=118.5 tau^2^=0.351 p<0.001). Egger’s method A=-4.128 p=0.003.

### Length of hospital stay

Among survivors, the length of hospital stay in the TCZ-group was a median of 15.3 days [range 12.4-19.4] vs. 14 days [range 9-20] in the control group. These differences were not statistically significant (p=0.953).

## DISCUSSION

Pending published evidence from RCTs, this SRMA focused on available real-world observational studies, revealing a beneficial effect of TCZ use in preventing mortality in hospitalized adults with COVID-19. However, the present results also showed a higher relative risk for ICU admission and the occurrence of secondary infections in such COVID-19 patients receiving TCZ.

To date, two existing SRMA have summarized current evidence on the beneficial and harmful effects of TCZ in COVID-19. The first by Lan SH Zhang et al., included 7 studies, with no conclusive evidence that TCZ would provide any additional benefit to patients with severe COVID-19 [77]. The second, registered in the medRxiv repository by Boregowda et al., included 16 studies, concluding that the addition of TCZ to SOC might reduce mortality in COVID-19 patients requiring hospitalization [78].

The present SRMA updated and expanded the revision to 57 observational studies. As expected, most included studies emerged recently and were performed in hospitals from high-income European countries and the US. The finding that most of the included patients were men in their 6-8th decades of life was consistent with what has already been described in the general population requiring hospital admission due to COVID-19 [79]. The vast majority of included studies used a single dose of 400-800 mg intravenous or 162-324 mg subcutaneous, and a few of them allowed a second or even a third dose in case of worsening. Interestingly, the present SRMA showed that TCZ was mainly prescribed as a second step to treat those patients at risk of transition to a more severe condition, after showing poor response to antiviral agents. Accordingly, in a notable proportion of included studies, TCZ was indicated in combination with corticosteroids. In this respect, in view of the RECOVERY trial [5] which reported significant benefit from the use of steroids in severe COVID-19, it is difficult to distinguish in depth the contribution of TCZ on the outcomes in such included patients receiving TCZ and steroids. On the other hand, in addition to TCZ, a very small proportion (<1%) of included patients from more recent studies also received remdesivir, as a proven antiviral against SARS-Cov-2.

In addition to concomitant drugs, other relevant factors should be taken into account when considering the impact of TCZ use on clinical outcomes in COVID-19. In this respect, in addition to the age and underlying comorbidities of included patients, one of the most important factors to be considered at the clinical level is the severity of the clinical-stage when indicating TCZ to treat the COVID-19 illness [6,79].

Unfortunately, the severity of patients could not be accurately inferred from the clinical, laboratory, or radiological parameters documented in the included studies of the present SRMA. Therefore, mortality was compared between those patients admitted to hospital wards and ICUs, in those studies in which the hospital site from where TCZ was administered was specified. Logically, patients in whom TCZ was indicated during ICU admission showed higher mortality in comparison to those treated in hospital wards. However, since the COVID-19 pandemic induced an unprecedented influx of patients into the ICUs, the particular emergency and resource availability of each hospital involved in the included studies most likely conditioned either the criteria when transferring patients from hospital wards to ICUs, or the ethical decisions related to the withdrawal of life support decisions.

Outside of RCTs, observational data from the included studies only reflected the clinical practice of physicians when indicating TCZ use. Consequently, the present results show that TCZ was mostly prescribed in those more seriously-ill patients presenting at a more advanced stage of COVID-19, with severe lung injury and systemic hyperinflammatory multi-organ failure. In this regard, it would be unfair to infer that the higher the use of TCZ and the average of concomitant corticosteroids, the higher the risk of ICU admission or death. Therefore, further RCTs on TCZ use in COVID-19 should clarify the interaction of confounding variables, it being crucial to know which patients are the best candidates to eventually receive TCZ as an immunomodulatory agent, as well as the beneficial and harmful effects of its use in the absence of or in combination with steroids.

Moreover, in the particular case of the ICU setting, most of the included studies showed insufficient data to appropriately assess the effects when TCZ was prescribed in critically-ill patients. However, since a majority of such seriously-ill patients with COVID-19 admitted to ICUs are submitted to mechanical ventilation and other multiple invasive procedures, and receive concomitant wide-spectrum antibiotics or steroid treatment, it is of great concern the well-known risk of TCZ favoring the occurrence of life-threatening secondary bacterial, viral or fungal opportunistic infections, as it has also been documented in the present SRMA in up to one-fifth of cases in the TCZ-group [53,80,81].

As mentioned, the first and major limitation of this SRMA is the lack of data from RCTs. In their absence, the present revision was based on observational studies; therefore, conclusions should be considered with caution. Moreover, a second limitation is the fact that most of the included studies were retrospective in nature. A third limitation is the heterogeneity regarding the study population (I^2^ index) and the potential risk of detected bias. Fourth, variations in criteria for prescribing TCZ may not be ruled out in the included studies, although most of them indicated TCZ use to treat those patients with severe COVID-19 with the systemic hyperinflammatory state. Fifth, important factors influencing the effect of TCZ on clinical outcomes such as the baseline characteristics of the patients included, the average time from symptoms onset to TCZ administration, the clinical severity of the disease at the time of TCZ administration, the doses and the form of administration used, the hospital site from where TCZ was indicated, or the use of concomitant drug regimens could not be evaluated in-depth, since they were not uniformly provided by the included studies. Sixth, in the vast majority of included studies, there is a lack of subgroup analyses according to age, sex or underlying conditions, concomitant treatments, the requirement of mechanical ventilation or ICU admission, and comparisons between ventilated and non-ventilated patients. Finally, there is a wide range in the median time of follow-up after TCZ administration, which hinders assessment of consistent improvement, late-onset adverse events, and real in-hospital mortality in those patients with prolonged evolution. Thus, some patients considered “survivors” in some included studies may have ended up dying.

In conclusion, pending evidence from RCTs, this systematic review provides updated and extended data from observational studies on the use of TCZ in COVID-19. The present results showed TCZ to be beneficial in reducing overall in-hospital mortality in adults with COVID-19, with an NNT to save one life of 20. These findings were more apparent in those non-critically-ill COVID-19 patients admitted to hospital wards, receiving TCZ at the early stage of hyperinflammatory response syndrome. By contrast, the TCZ-group was at higher risk for secondary infections, especially in those patients admitted to ICU. Notwithstanding these results, conclusions should be considered as weak evidence since they are based on observational studies, most of them retrospective. However, these findings may help physicians and researchers to optimize strategies towards precision medicine when designing further RCTs focused on the use of TCZ in COVID-19.

## Data Availability

The datasets generated during and/or analyzed during the current study are available from the corresponding author on reasonable request.

## ACKNOWLEDGMENTS

We dedicate this work to the memory of those patients worldwide who have not survived COVID-19. We thank CERCA Programme/Generalitat de Catalunya for institutional support.

## AUTHORS’ CONTRIBUTION

Study conception and design: M.R-R., X.C.

Acquisition, analyses and interpretation of data: M.R-R., J.M.M-L, A.M., N.A.H., J.R., X.C. Manuscript draft and critical revision: M.R-R., J.R., X.C.

